# Comparison of the Diagnostic Performance from Patient’s Medical History and Imaging Findings between GPT-4 based ChatGPT and Radiologists in Challenging Neuroradiology Cases

**DOI:** 10.1101/2023.08.28.23294607

**Authors:** Daisuke Horiuchi, Hiroyuki Tatekawa, Tatsushi Oura, Satoshi Oue, Shannon L Walston, Hirotaka Takita, Shu Matsushita, Yasuhito Mitsuyama, Taro Shimono, Yukio Miki, Daiju Ueda

**Affiliations:** Department of Diagnostic and Interventional Radiology, Graduate School of Medicine, Osaka Metropolitan University, Osaka, Japan; Smart Life Science Lab, Center for Health Science Innovation, Osaka Metropolitan University, Osaka, Japan

**Keywords:** Artificial intelligence, Chat Generative Pre-trained Transformer (ChatGPT), Generative Pre-trained Transformer (GPT)-4, Large language models

## Abstract

**Purpose:** To compare the diagnostic performance between Chat Generative Pre-trained Transformer (ChatGPT), based on the GPT-4 architecture, and radiologists from patient’s medical history and imaging findings in challenging neuroradiology cases.

**Methods:** We collected 30 consecutive “Freiburg Neuropathology Case Conference” cases from the journal Clinical Neuroradiology between March 2016 and June 2023. GPT-4 based ChatGPT generated diagnoses from the patient’s provided medical history and imaging findings for each case, and the diagnostic accuracy rate was determined based on the published ground truth. Three radiologists with different levels of experience (2, 4, and 7 years of experience, respectively) independently reviewed all the cases based on the patient’s provided medical history and imaging findings, and the diagnostic accuracy rates were evaluated. The Chi-square tests were performed to compare the diagnostic accuracy rates between ChatGPT and each radiologist.

**Results:** ChatGPT achieved an accuracy rate of 23% (7/30 cases). Radiologists achieved the following accuracy rates: a junior radiology resident had 27% (8/30) accuracy, a senior radiology resident had 30% (9/30) accuracy, and a board-certified radiologist had 47% (14/30) accuracy. ChatGPT’s diagnostic accuracy rate was lower than that of each radiologist, although the difference was not significant (*p* = 0.99, 0.77, and 0.10, respectively).

**Conclusion:** The diagnostic performance of GPT-4 based ChatGPT did not reach the performance level of either junior/senior radiology residents or board-certified radiologists in challenging neuroradiology cases. While ChatGPT holds great promise in the field of neuroradiology, radiologists should be aware of its current performance and limitations for optimal utilization.

## Introduction

Chat Generative Pre-trained Transformer (ChatGPT) is a cutting-edge large language model developed by the OpenAI company [1]. ChatGPT, based on the GPT-4 architecture, has remarkable capabilities in understanding natural languages and generating human-like text responses across a wide variety of topics [2-4]. ChatGPT holds the promise to revolutionize many industries, and professionals in various fields are considering its implementation to enhance efficiency and support decision-making processes [5].

Artificial intelligence has already been applied to clinical applications in the field of radiology, showing remarkable benefits [6-8]. ChatGPT holds the potential to be a valuable tool in radiology, and several initial applications of ChatGPT have been reported [9-18]. GPT-3.5 based ChatGPT almost passed a text-based radiology examination without specific radiology training, and the subsequent GPT-4 based ChatGPT has passed the examination, surpassing the performance of its predecessor [19, 20]. Considering its potential for clinical applications in radiology, radiologists need to be aware of ChatGPT’s current performance and limitations for optimal utilization.

Diagnostic neuroradiology is a complex field that requires specialized expertise to interpret diverse imaging findings associated with various diseases [21]. Radiologists may benefit from the assistance provided by ChatGPT, especially in diagnosing complex and challenging cases. Recent studies have reported the diagnostic performance of GPT-4 based ChatGPT in the field of radiology [9, 10]; however, ChatGPT’s diagnostic performance in challenging neuroradiology cases and its comparison with radiologists’ diagnostic performance have not yet been investigated and remain unclear. The journal Clinical Neuroradiology presents diagnostic cases in the “Freiburg Neuropathology Case Conference” section that are both educational and interesting, as well as complex and challenging for clinicians. By comparing the diagnostic performance of ChatGPT and radiologists in these cases, we can gain valuable insights into the capabilities of ChatGPT in neuroradiology.

This study aimed to compare the diagnostic performance, based on patient’s medical history and imaging findings, between GPT-4 based ChatGPT and radiologists in challenging neuroradiology cases using the “Freiburg Neuropathology Case Conference” cases published in Clinical Neuroradiology.

## Methods

### Study design

In this study, we input the patient’s medical history and imaging findings into ChatGPT, which generated differential and final diagnoses. We utilized imaging findings instead of the images themselves, as the current version of ChatGPT could not directly process images. The diagnostic performance of ChatGPT was evaluated by assessing the accuracy rate of the ChatGPT’s diagnosis. The study design adhered to the Standards for Reporting Diagnostic Accuracy Studies statement [22]. Ethics committee approval was not required since this study utilized only published cases.

### Data collection

The journal Clinical Neuroradiology publishes diagnostic cases in the “Freiburg Neuropathology Case Conference” section, with one case being published per issue. The 2016 World Health Organization (WHO) Classification of Tumours of the Central Nervous System (CNS) introduced a molecular tumor classification into the diagnostic framework of CNS tumors, and the latest 2021 WHO Classification of Tumours of the CNS built upon the molecular approach, adding more molecular features and updating pathologic diagnoses [23]. Given the paradigm shift of the WHO Classification of Tumours of the CNS in 2016, we included the “Freiburg Neuropathology Case Conference” cases from 2016 onward and collected 30 consecutive cases from March 2016 (volume 26, issue 1) to June 2023 (volume 33, issue 2). We collected the patient’s medical history from the “Case Report” section, the imaging findings from the “Imaging” section, and the diagnosis (actual ground truth) from the “Diagnosis” section of each case. The “Case Report” section contained the descriptions of biopsy/surgical findings and postoperative clinical course; thus, we excluded these descriptions from the patient’s medical history. Fig. 1 shows the data collection flowchart.

**Fig. 1.**
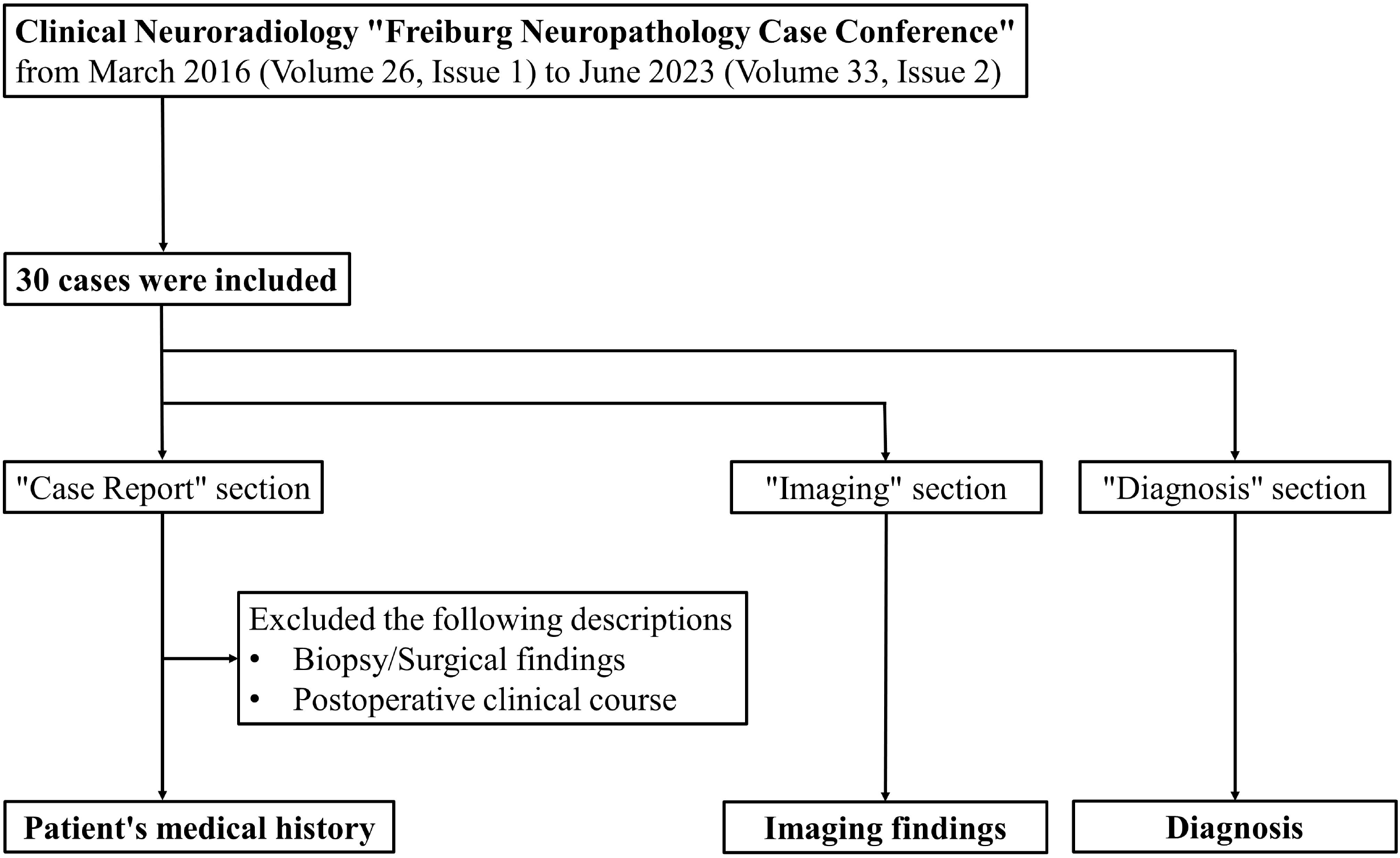
Data collection flowchart

### Input/output procedure for ChatGPT and Output evaluation

We initially entered the following prompt as the task into ChatGPT based on the GPT-4 architecture (May 24 version; OpenAI, California, USA; https://chat.openai.com/): *As a physician, I plan to utilize you for research purposes. Assuming you are a hypothetical physician, please walk me through the process from differential diagnosis to the most likely disease step by step, based on the patient’s information I am about to present. Please list three possible differential diagnoses in order of likelihood*. Subsequently, we input the patient’s medical history and imaging findings and obtained the output from ChatGPT for each case (an illustrative example is presented in Fig. 2). We started a new ChatGPT session for each case and input both the prompt and the patient’s medical history/imaging findings to prevent any potential influence of previous answers on ChatGPT’s output. These processes were conducted once for each case between June 8 and June 9, 2023.

**Fig. 2.**
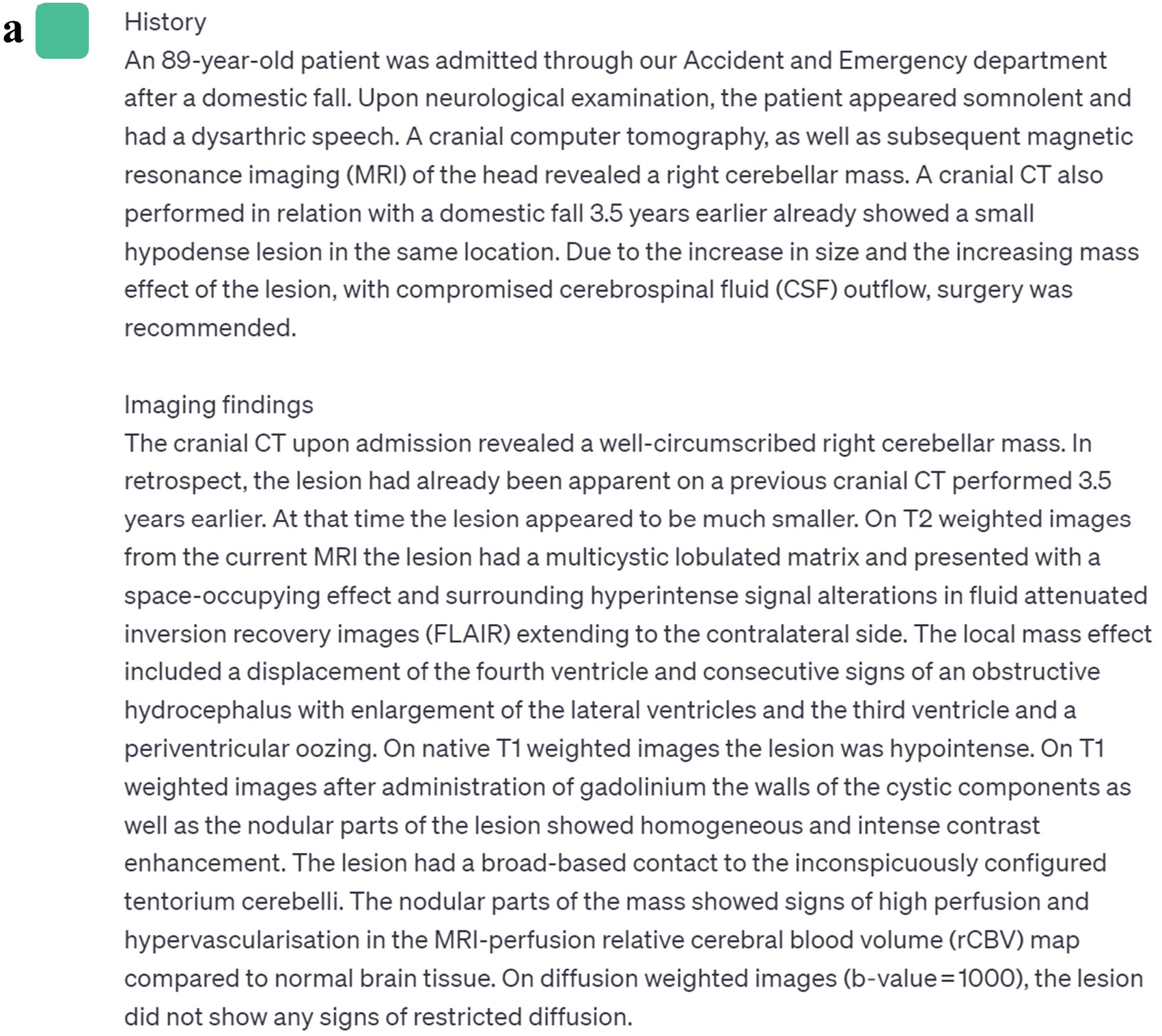
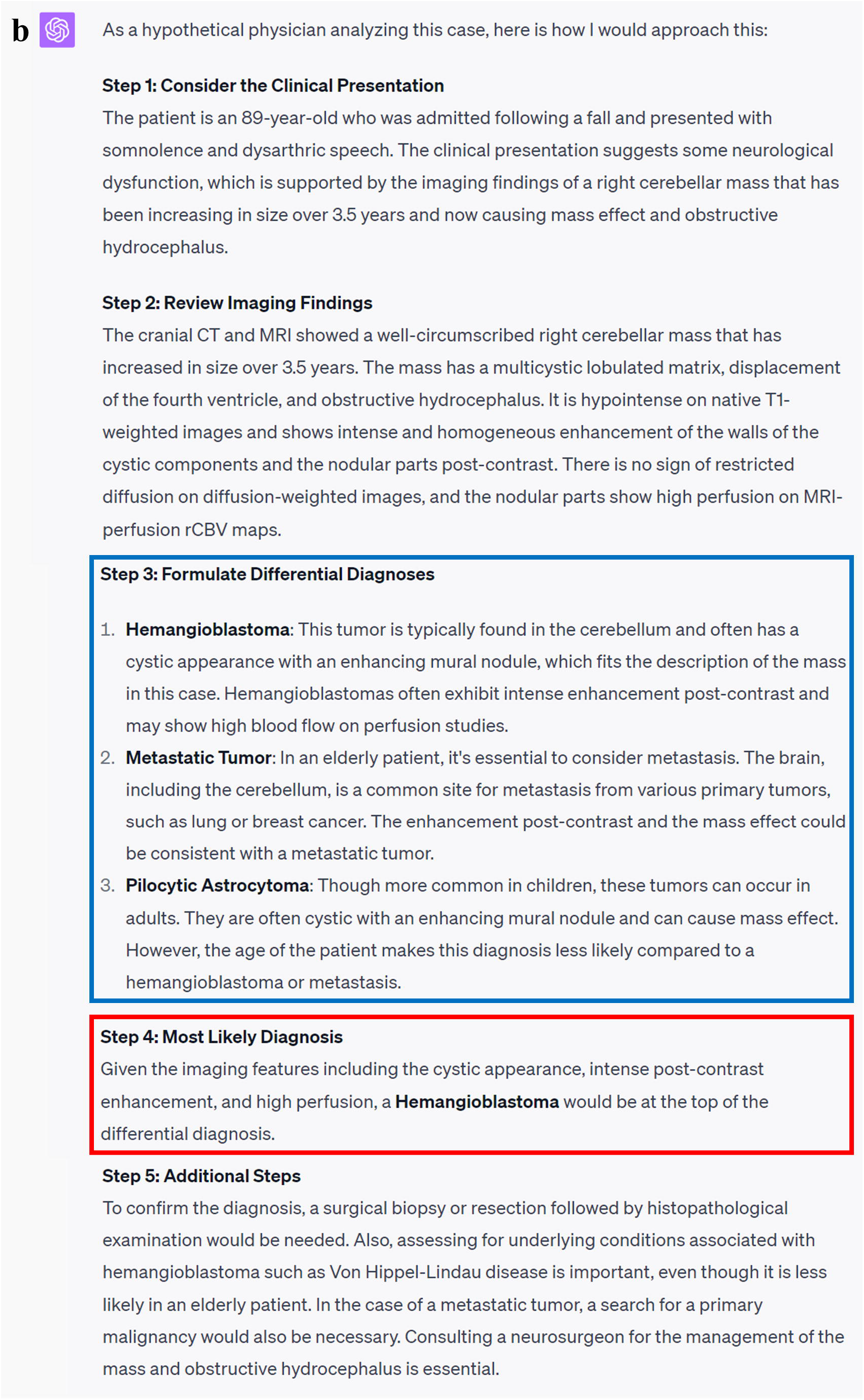
An illustrative example of the input to and output from ChatGPT. **a** Input texts (patient’s medical history and imaging findings) to ChatGPT. **b** Output texts generated by ChatGPT. The differential diagnoses are highlighted in the blue area, and the final diagnosis is highlighted in the red area. In this case [29], the final diagnosis generated by ChatGPT was correct

The output generated by ChatGPT consisted of three differential diagnoses and one final diagnosis chosen from them. Two board-certified radiologists (13 years of experience [H.T.]; 7 years of experience [D.H.]) determined whether the differential diagnoses and final diagnosis generated by ChatGPT aligned with the actual ground truth. If there were any discrepancies, a final decision was made by consensus.

### Radiologists’ interpretation

All 30 cases from the “Freiburg Neuropathology Case Conference” were independently reviewed by three radiologists with different levels of experience: one junior radiology resident (Reader 1 [S.O.]; 2 years of experience in radiology), one senior radiology resident (Reader 2 [T.O.]; 4 years of experience in radiology, including 1 year of training in neuroradiology), and one board-certified radiologist (Reader 3 [D.H.]; 7 years of experience in radiology, including 4 years of training in neuroradiology). Each radiologist conducted their diagnoses based on the “Case Report” (excluding the descriptions of biopsy/surgical findings and postoperative clinical course) and the “Imaging” sections (both the description of the imaging findings and the images themselves). They provided three differential diagnoses and one final diagnosis chosen from them for each case. All radiologists were blinded to the differential and final diagnoses generated by ChatGPT, as well as the actual ground truth. The accuracy rates of these diagnoses were considered as the radiologists’ diagnostic performance.

### Statistical analysis

Statistical analyses were performed with R software (version 4.0.2, 2020; R Foundation for Statistical Computing, Vienna, Austria; http://www.r-project.org/). As the current GPT-4 based ChatGPT has been trained on data available up to September 2021 [1], the cases published until September 2021 had potential for bias. Thus, we categorized the cases into two groups: those with publication dates through September 2021 and those from October 2021 onward. We performed pairwise Fisher’s exact tests to compare the diagnostic accuracy rates of the final diagnosis and the differential diagnoses between the two groups. Additionally, we performed the Chi-square tests to compare the diagnostic accuracy rates of the final diagnosis and differential diagnoses between ChatGPT and each radiologist. Adjustment for multiplicity was not performed because this was an exploratory study. A two-sided *p* value < 0.05 was considered statistically significant.

## Results

The 30 cases from the “Freiburg Neuropathology Case Conference” cases consisted of 27 cases of neoplastic diseases and 3 cases of non-neoplastic diseases. ChatGPT successfully generated one final diagnosis and three differential diagnoses for each case and exhibited a final diagnostic accuracy of 23% (7/30 cases) and a differential diagnostic accuracy of 40% (12/30 cases) (Table 1). The final diagnostic accuracy rates were 17% (4/23 cases) for the cases published through September 2021 and 43% (3/7 cases) for those from October 2021 onward, while the differential diagnostic accuracy rates were 39% (9/23 cases) for the cases through September 2021 and 43% (3/7 cases) for those from October 2021 onward. No significant difference was observed in either the final or differential diagnostic accuracy rates between the two periods (*p* = 0.31 and 0.99, respectively).

**Table 1.** ChatGPT’s diagnostic accuracy.

|  | Correct answer (accuracy rate [%]) |  |
| --- | --- | --- |
|  | Final diagnosis | Differential diagnosis |
| Overall diagnostic accuracy | 7/30 (23%) | 12/30 (40%) |
| Etiology |  |  |
| Neoplastic disease | 6/27 (22%) | 10/27 (37%) |
| Non-neoplastic disease | 1/3 (33%) | 2/3 (67%) |
| Publication date |  |  |
| Until September 2021 | 4/23 (17%) | 9/23 (39%) |
| From October 2021 onward | 3/7 (43%) | 3/7 (43%) |

Regarding the radiologists’ interpretations, the accuracy rates for the final and differential diagnoses were as follows: Reader 1 (junior radiology resident) achieved accuracy rates of 27% (8/30) and 47% (14/30), Reader 2 (senior radiology resident) achieved accuracy rates of 30% (9/30) and 63% (19/30), and Reader 3 (board-certified radiologist) achieved accuracy rates of 47% (14/30) and 70% (21/30). Among the three radiologists, those with more years of experience demonstrated higher diagnostic accuracy rates in both the final and differential diagnoses.

When comparing ChatGPT and radiologists, ChatGPT’s diagnostic accuracy rates for the final and differential diagnoses were lower than those of each radiologist. Regarding the final diagnostic accuracy rates, no significant difference was observed between ChatGPT and each radiologist (*p* = 0.99, 0.77, and 0.10, respectively). As for the differential diagnostic accuracy rates, no significant difference was observed between ChatGPT and Reader 1 or Reader 2 (*p* = 0.79 and 0.12, respectively), while Reader 3 showed a significantly higher accuracy rate compared to ChatGPT (*p* = 0.04) (Table 2).

**Table 2.** Comparison of the diagnostic accuracy between ChatGPT and radiologists.

|  | Correct answer (accuracy rate [%]) |  |  |  |
| --- | --- | --- | --- | --- |
|  | Final diagnosis | *p* value * | Differential diagnosis | *p* value * |
| GPT-4 based ChatGPT | 7/30 (23%) |  | 12/30 (40%) |  |
| Reader 1 (Junior radiology resident) | 8/30 (27%) | 0.99 | 14/30 (47%) | 0.79 |
| Reader 2 (Senior radiology resident) | 9/30 (30%) | 0.77 | 19/30 (63%) | 0.12 |
| Reader 3 (Board-certified radiologist) | 14/30 (47%) | 0.10 | 21/30 (70%) | 0.04** |

GPT; Generative Pre-trained Transformer
\* The Chi-square tests are performed to compare the accuracy rates between ChatGPT and each radiologist
\*\* $p < 0.05$

## Discussion

This study compared the diagnostic performance, based on patient’s medical history and imaging findings, between GPT-4 based ChatGPT and radiologists with various levels of experience in challenging diagnostic cases in neuroradiology. GPT-4 based ChatGPT achieved a final diagnostic accuracy of 23% (7/30 cases) and a differential diagnostic accuracy of 40% (12/30 cases) for the “Freiburg Neuropathology Case Conference” cases published in Clinical Neuroradiology between March 2016 and June 2023. No significant difference was observed in the diagnostic accuracy rates of ChatGPT between the cases published until September 2021 and those from October 2021 onward. ChatGPT’s final and differential diagnostic accuracy rates were lower than those of a junior radiology resident, a senior radiology resident, and a board-certified radiologist, although not significantly so. Only the board-certified radiologist had a significantly higher differential diagnostic accuracy compared to ChatGPT.

To the best of our knowledge, this study is the first to compare the diagnostic performance of GPT-4 based ChatGPT and radiologists in challenging neuroradiology cases. Although a previous study has reported the diagnostic performance of GPT-4 based ChatGPT from patient’s medical history and imaging findings in general radiology [10], no study has evaluated and compared the diagnostic performance of ChatGPT and radiologists on challenging neuroradiology cases. This study found that the diagnostic performance of GPT-4 based ChatGPT did not reach the performance level of either junior/senior radiology residents or board-certified radiologists in challenging neuroradiology cases.

ChatGPT has the potential to improve the clinical workflow in radiology [24, 25]. Several studies have reported that ChatGPT offers valuable assistance to radiologists in various tasks, including supporting diagnosis/decision-making, determining imaging protocols, generating/simplifying radiology reports, writing medical publications, and providing patient education [9-18]. With the advancement of medical imaging technologies and the overutilization of imaging examinations, the workload for radiologists has increased, thereby contributing to diagnostic errors in neuroradiology [26, 27]. Integrating ChatGPT as a diagnostic tool in clinical practice is expected to save radiologists’ interpretation time and reduce their workload [9, 10], potentially leading to a decrease in diagnostic errors and improved patient outcomes.

While ChatGPT has the potential to revitalize the field of neuroradiology, radiologists need to recognize its limitations and exercise caution when integrating ChatGPT into clinical practice. This study demonstrated that the diagnostic performance of GPT-4 based ChatGPT did not reach the performance level of either junior/senior radiology residents or board-certified radiologists in challenging neuroradiology cases. Radiologists may need the diagnostic assistance provided by ChatGPT, especially in complex and challenging cases. However, our results indicated that ChatGPT’s diagnostic performance is inadequate in challenging neuroradiology cases, and the current ChatGPT cannot fully replace the expertise of radiologists. The majority of cases in this study were neoplastic diseases, and the wide variety of histopathological types and imaging findings associated with these neoplastic diseases may have contributed to ChatGPT’s insufficient diagnostic accuracy [23, 28]. In addition, radiologists should be aware that the output generated by ChatGPT may not fully correspond to the 2021 WHO classification of CNS tumors [23], given that the current ChatGPT has been trained on data up to September 2021 [1]. Furthermore, the current ChatGPT’s diagnostic performance in clinical practice should be considered dependent on the radiologist’s ability, as it cannot directly process images and relies on the inputs of imaging findings provided by radiologists. The development of ChatGPT-based algorithms may improve these limitations, thus radiologists need to be familiar with these rapidly evolving technologies for optimal utilization.

This study had several limitations. First, this study included a relatively small sample size, which limits the statistical power of the analyses. Second, ChatGPT’s diagnostic performance was evaluated in a controlled environment using the “Freiburg Neuropathology Case Conference” cases, which may not accurately reflect the complexities and challenges of real-world clinical practice. Third, this study utilized the “Freiburg Neuropathology Case Conference” cases in Clinical Neuroradiology as challenging cases in the field of neuroradiology; however, the definition of challenging neuroradiology cases may be inherently subjective.

Further studies are required to explore various types of challenging diagnostic cases in neuroradiology. Finally, since the majority of cases in this study were neoplastic diseases, the comparison of diagnostic performance between ChatGPT and radiologists may be inadequate for non-neoplastic diseases.

## Conclusion

This study demonstrated that the diagnostic performance of GPT-4 based ChatGPT did not reach the performance level of either junior/senior radiology residents or board-certified radiologists in challenging neuroradiology cases. These findings indicate that the current version of ChatGPT cannot fully replace the expertise of radiologists. While ChatGPT holds great promise in the field of neuroradiology, radiologists should be aware of its current performance and limitations for optimal utilization. Further improvements, such as fine-tuning the GPT-4 model to achieve higher performance in radiology tasks, could be future research.

## Data Availability

All data produced in the present work are contained in the manuscript.

## Notes

### Competing Interest Statement

The authors have declared no competing interest.

### Funding Statement

This study did not receive any funding

